# How side effects can improve treatment efficacy: a randomized trial

**DOI:** 10.1101/2023.11.22.23298877

**Authors:** Lieven A. Schenk, Tahmine Fadai, Christian Büchel

## Abstract

While treatment side effects may adversely impact patients, they could also potentially function as indicators for effective treatment. In this study, we investigated whether and how side effects can trigger positive treatment expectations and enhance treatment outcomes.

In this preregistered trial (DRKS00026648), 77 healthy participants were made to believe that they will receive fentanyl nasal sprays before receiving thermal pain in a controlled experimental setting. However, nasal sprays did not contain fentanyl, rather they either contained capsaicin to induce a side effect (mild burning sensation) or saline (control). Following the initial phase, participants were randomized to two groups and underwent functional magnetic resonance imaging (fMRI). One group continued to believe that the nasal sprays could contain fentanyl while the other group was explicitly informed that no fentanyl was included. This allowed for the independent manipulation of the side effects and the expectation of pain relief.

Our results revealed that nasal sprays with a side effect lead to lower pain than control nasal sprays without side effects. The influence of side effects on pain was dependent on individual beliefs about how side effects are related to treatment outcome, as well as on expectations about received treatment. FMRI data indicated an involvement of the descending pain modulatory system including the anterior cingulate cortex and the periaqueductal gray during pain after experiencing a nasal spray with side effects.

In summary, our data show that mild side effects can serve as a signal for effective treatment thereby influencing treatment expectations and outcomes, which is mediated by the descending pain modulatory system. Using these mechanisms in clinical practice could provide an efficient way to optimize treatment outcome. In addition, our results indicate an important confound in clinical trials, where a treatment (with potential side effects) is compared to placebo.

## Introduction

Common thinking in modern medicine posits that ideal treatments should have no side effects, because they can cause discomfort, suffering and treatment discontinuation. In addition, the mere expectation of side effects increases the likelihood of side effects^1^. Consequently, it has been suggested to not only minimize side effects, but also to carefully disclose information regarding potential side effects to avoid these nocebo effects^2^.

Here we challenge this view and ask whether some side effects could actually lead to better treatment outcomes. This idea is motivated by the observation that side effects themselves can contribute to treatment expectations^3^. Our hypothesis posits that even mild side effects can be indirect indicators of treatment potency (e.g. side effects are unavoidable with a powerful drug), which can lead to positive treatment expectations. These treatment expectations are the basis for non-specific therapeutic effects (i.e. placebo effect) that have substantial impact on treatment outcomes^4–6^.

Support for this hypothesis comes from research on active placebos, i.e. pharmacological agents that have a noticeable effect on the patient but not on the primary symptoms. A previous study on pain suggested that active placebos can indeed lead to larger placebo effects than inert placebos^7^. This idea also resonates with the observation that general practitioners prescribed more impure (i.e. active) placebos than inert placebos^8^.

The question of whether side effects can influence treatment outcome is also relevant for randomized clinical trials, as most studies compare active pharmacological interventions with inert placebos^9^. If the experience of side effects can indirectly boost treatment outcome, a clinical trial might overestimate the beneficial effect of active treatments that have an easily identifiable side effect profile.

To address these questions, we designed a multistep experiment to investigate how side effects influence treatment efficacy on the psychological and neural level in a large sample of healthy volunteers using fMRI (Figure 1). The study was done in a controlled experimental placebo paradigm to exclude hidden pharmacological effects and isolate psychological and neural mechanisms, therefore it is not related to any drug-specific pharmacological effects and can be generalized to other treatments. According to our preregistration (https://drks.de/search/de/trial/DRKS00026648), we hypothesized that side effects act as a cue for effective treatment and influence pain and placebo effects by augmenting expectation mechanisms. With respect to neuronal effects we hypothesized that side effects would recruit the descending pain modulatory system and in particular that the coupling between the rostral ACC and the PAG is modulated.

**Figure 1:**
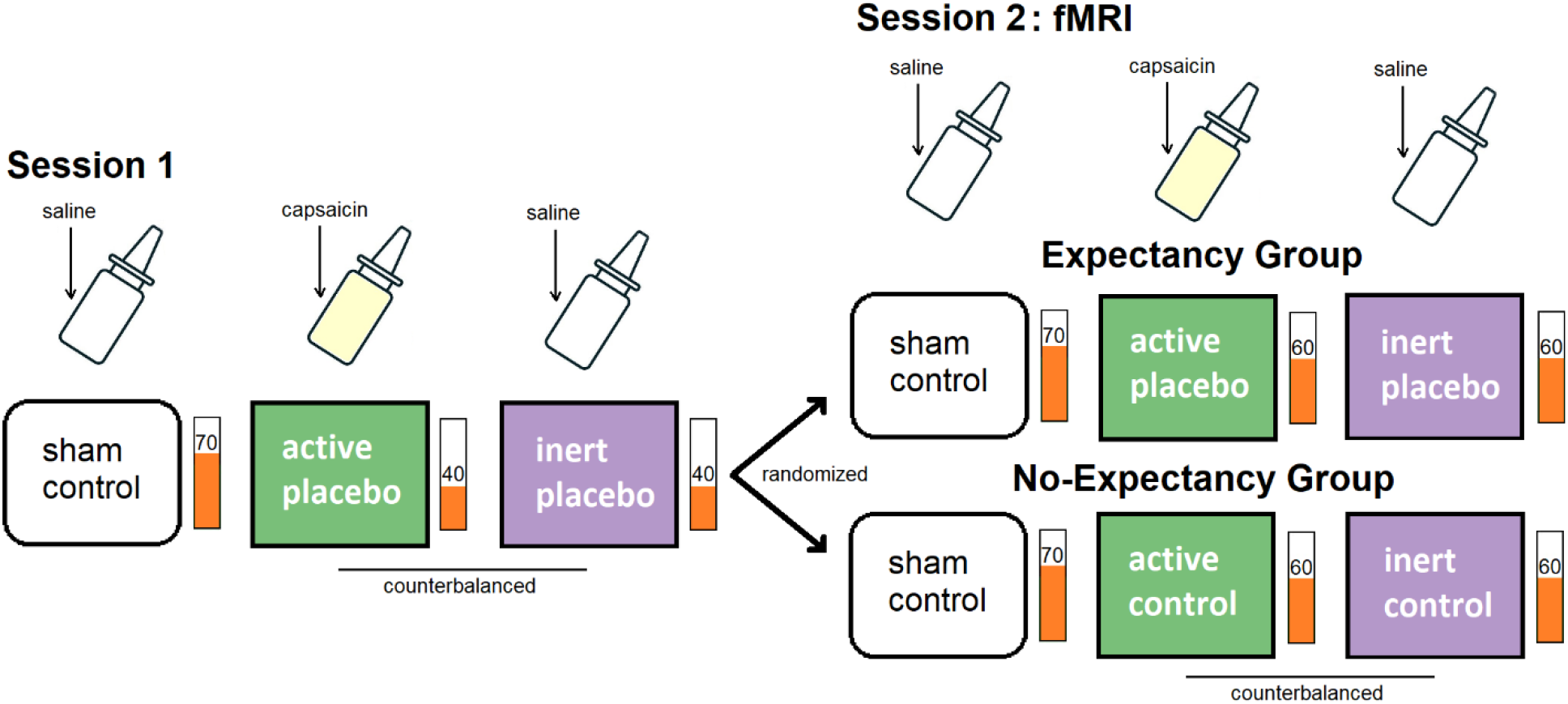
Experimental Design. In the first session, participants received three different nasal sprays (in three separate runs) with the information that each of them could contain the potent pain killer fentanyl. Unbeknownst to the participants, none of the nasal sprays contained any active pain medication, but one of the nasal sprays contained a small dose of capsaicin, causing a mild, but clearly perceptible burning-like sensation in the nose. After the application of the nasal sprays, a series of thermal pain stimuli was applied (corresponding to previously calibrated levels of 40, 60 or 70 on a visual analogue scale (VAS) from 0 to 100) and participants rated their pain on the VAS. Pain was reduced in the placebo conditions to mimic a treatment benefit. In a similar second session, the same procedure was repeated during fMRI to investigate the neural mechanisms of how side effects can modulate treatment efficacy. To test for the role of expectation of treatment benefit, the sample was randomized into two groups before the second session. The expectation group believed that the experiment will be repeated and continued to expect that any pain decreases were due to the pain relieving fentanyl nasal spray, whereas the no-expectation group was explicitly debriefed that no fentanyl was present in any nasal spray and that all differences in pain were due to differences in the applied temperature. Therefore, side effects and expectation of treatment benefit were manipulated in an independent fashion, which allowed us to not only investigate the role of side effects and expectation, but also the interaction between both factors.

## Materials and Methods

### Participants

104 healthy participants were enrolled in this study. Nine participants were excluded due to problems with the nasal spray, one due to a technical problem and two due to floor effects of applied pain (mean ratings < VAS10). 15 participants were excluded after performing the preregistered manipulation check (see Supplementary Information). 77 participants were included in the data analysis (42 expectancy group (age: 24.3±3.8 [18-38], 12 male); 35 no-expectancy group (age: 24.8±5.4 [18-43], 14 male)). Participants were confirmed to be healthy with an in-person interview with a medical doctor during the initial visit (see Supplementary Information). All participants gave written consent according the Declaration of Helsinki and the study was approved by the Ethics Committee of the Hamburg Medical Association.

### Experimental paradigm

The experiment consisted of 3 visits. During the initial visit, participants were informed about the research, checked for eligibility, and signed consent forms. Participants underwent a medical counseling session were they received information that the aim of the study was to investigate the neural processes associated with fentanyl nasal spray, a powerful analgesic drug used in the treatment of cancer pain. They were informed about the medication’s purpose and potential side effects of fentanyl, which included a burning sensation in the nose. Afterwards, basic vital information (height, weight, blood pressure) was assessed and a drug screening was performed. Participants then completed a series of questionnaires, including questions about their belief that side effects indicate a more potent treatment.

During the experimental visit, critical instructions were repeated and a heat pain sensitivity assessment was performed. Heat pain stimuli were applied using a thermode (PATHWAY System, Medoc, Ramat Yishai, Israel). Participants were then instructed how to use a visual analogue scale (VAS) to rate their pain (ranging from 0=no pain to 100=maximum tolerable pain). Finally, participants rated the intensity of a series of painful heat stimuli to establish temperature levels corresponding to individual pain at VAS 40, 60 and 70.

During the experiment, participants believed that they receive three nasal sprays in three experimental runs, each with a 50% chance of containing fentanyl, and that this procedure will be repeated in the MR scanner as well as 7±1 days later (Figure 1). Unbeknownst to the participants, none of the nasal sprays contained any fentanyl. However, one of the three nasal sprays (applied second or last) contained a small dose of capsaicin (0,15 μg/puff), which causes a burning sensation in the nose. After the application of each nasal spray, participants had to rate several items regarding potential side effects on a 4-point scale (“none at all”, “minimally”, “a bit”, “clearly”), including experiencing a burning sensation in the nose. Before each run, the thermode was moved to avoid sensitization or habituation and three warm-up stimuli were applied (20s). Each run consisted of 24 trials, and each trial consisted of anticipation (1.5s-2.5s), pain stimulation (6.5s with 4s pain plateau), VAS pain rating (8s) and a variable inter-trial-interval (7-9s).

The difference between the runs was the potential side effect experience (capsaicin or no capsaicin) and the applied temperature (VAS 40, 60 or 70; Figure 1). The first nasal spray always corresponded to the sham control condition. During this condition, the nasal spray did not cause any side effects (saline, no capsaicin) and pain corresponded to VAS 70. During the following two runs, participants received reduced pain corresponding to VAS 40 with either a saline nasal spray with capsaicin (active placebo) or without capsaicin (inert placebo). Stimulation temperature was reduced in both placebo conditions to mimic a treatment benefit compared to the previous sham control run. Both runs were the conditions of interest and the order was randomized across participants and concealed to participants and experimenters.. Afterwards, participants answered questions regarding their expectation of what they received in the previous runs and on how sure they were about their answer.

After the three runs, but before the MRI measurements, volunteers were randomized into an expectation and a no-expectation group. The randomized group allocation for all participants was performed at the start of data collection using a custom MATLAB script. The no-expectation group was debriefed to eliminate their expectation of pain relief, similar to previous experiments^10^. They were told that no fentanyl was present in any of the nasal sprays and that the pain was surreptitiously reduced during the second and third run. The expectation group was not informed and simply received the instruction that the same procedure will be repeated in the MR scanner. During MR scanning, participants completed the same paradigm as before, with the exception that during the second and third run they received pain corresponding to VAS 60. Participants were taken out of the scanner between runs and moved into a seating position to apply the nasal sprays, so that they could see that the same nasal sprays were applied. Finally, participants were invited for a follow-up visit (see Supplementary Information). At the end of the experiment, the expectation group was also debriefed. To account for the deceptive component, we reinstated participants’ autonomy by explicitly asking them whether they would like to withdraw their data at this stage. However, none of the participants withheld their approval.

### Behavioral data analysis

Behavioral data analysis was performed using SPSS 27 (IBM, Armonk, USA). Pain rating data were analyzed using repeated measures ANOVAs with condition (within) and group (between) as predictors and pain ratings as dependent variables, separate for each phase. For within-subject contrasts, paired-T-test and Wilcoxon Signed-Ranks Test were used, depending on scale and normal distribution. Multiple comparisons were Bonferroni-corrected. For mediation analysis, differences (active – inert) were calculated for side effects, treatment expectations (with guess treatment=2, no guess=1, guess control=0) and pain rating. The belief of how side effects influence treatment was assessed with their agreement (1-5) with the question: “stronger treatments have more side effects”. Mediation analysis was then performed with PROCESS in SPSS (model 8) with 5000 bootstrap samples to test for significance. All effects were considered significant at P<0.05 (two tailed).

### fMRI data analysis

fMRI data preprocessing and statistical analyses were performed using SPM12 (Wellcome Department of Imaging Neuroscience, London, UK). Data preprocessing consisted of slice timing, motion correction and coregistration of the functional images to the T1 anatomical scan. Finally, the images were spatially normalized using DARTEL (based on the IXI555 template from the CAT12 toolbox, http://dbm.neuro.uni-jena.de/cat/) and smoothed using a 6-mm (FWHM) isotropic Gaussian kernel.

We performed a first level analysis using a general linear model in SPM12. Each regressor was modeled by boxcar functions convolved with a canonical hemodynamic response function (HRF). For each run, we included regressors for cue, pain and pain rating. T-contrasts of interest were then calculated. T-tests between conditions of interest were used to test for significance. To maximize power, we employed a ROI approach according to our preregistration and selected ROIs based on previous meta-analyses: rostral ACC (interaction/inert contrast, 10mm)^11^; DLPFC (interaction/inert contrast, 10mm)^11^; insula (inert contrast, 8mm)^11^; S2 (inert contrast, 8mm)^11^ and PAG (inert contrast/connectivity analysis, 4mm)^12^. For medial regions (rostral ACC and the PAG), one ROI (with x=0) was used. For each contrast of interest, all ROIs were combined into a single mask to avoid inflation of Type-I Error. Results were considered significant at p<0.05 FWE corrected.

To test for a modulation of coupling between the rACC and the PAG, we performed a Psycho-Physiological Interaction (PPI) analysis^13^. We extracted the time series within a 3 mm sphere around the peak activation of the rACC from the interaction contrast [-6 33 -1.5] during the pain phase. Then we calculated the PPI interaction term as the time series multiplied by the psychological predictor (pain vs no pain). All three regressors were subsequently included in a new first level analysis. After model estimation, t-contrasts of interest were calculated for the PPI interaction, and PAG modulation was investigated with a ROIs of 4mm radius^12^.

For illustration purposes, all statistical maps use a significance threshold of p < 0.005 uncorrected and were overlaid on the mean structural image of all participants. All activations are reported using x, y, z coordinates in MNI (Montreal Neurological Institute) standard space.

## Results

### Side effects lead to improved pain relief and are mediated by treatment expectations

For the first session, we established that participants experienced more side effects after active placebo (p<0.001; F(1,75)=1138; Figure 2A). No interaction (p=0.65) or main effect of group (p=0.89) was observed. While all participants expected pain relief from the nasal sprays, we observed that pain ratings were lower after active placebo as compared to inert placebo (p=0.002; F(1,75)=10.7; VAS 26.2±2.0 vs 30.2±2.0; Figure 2B; Figure S1). As expected, there was no interaction (p=0.60) and no main effect of group (p=0.33). This clearly shows that the experience of side effects caused more pronounced pain relief.

**Figure 2:**
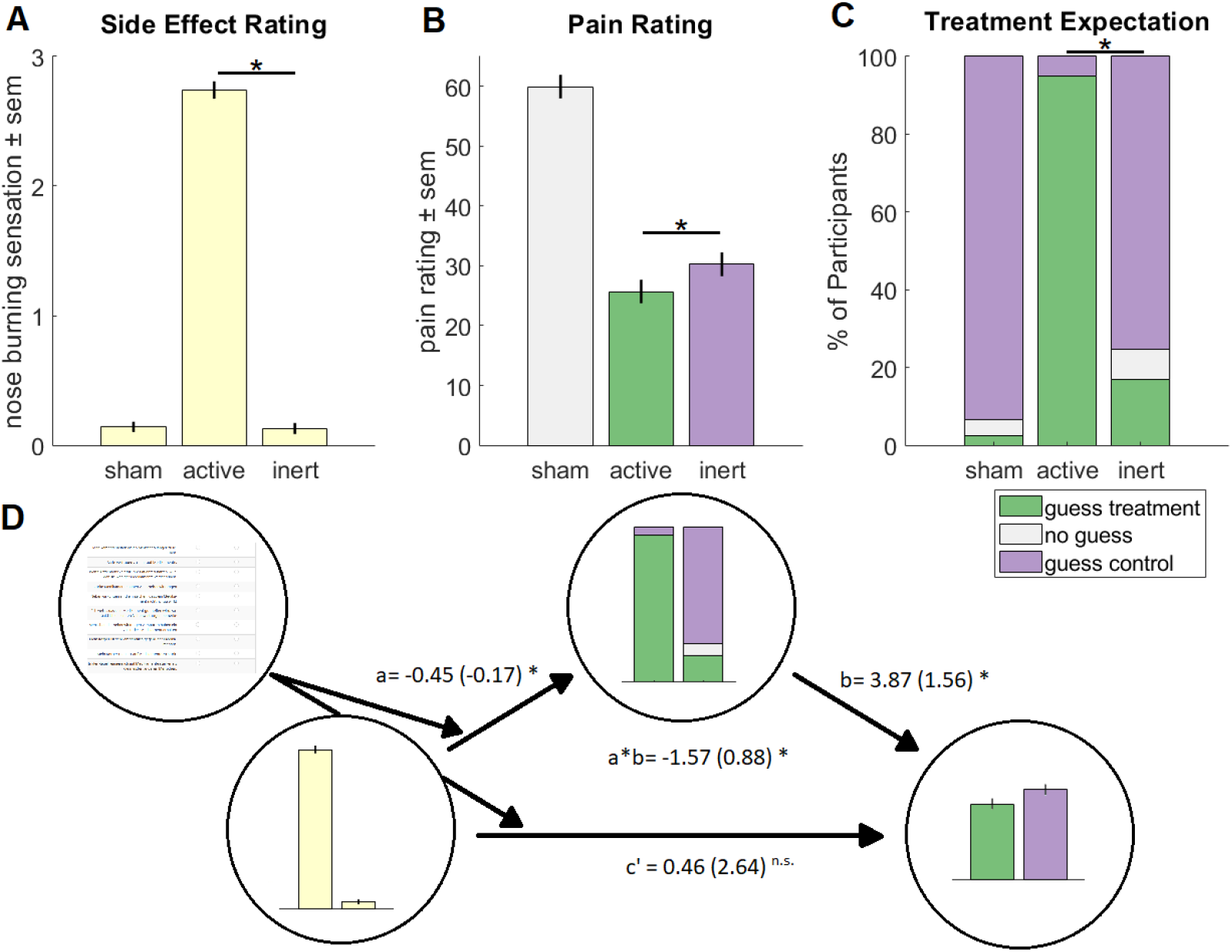
Session 1 Results. Side effects lead to larger pain relief and are mediated by treatment expectations. **A)** Participants experienced more side effects after active placebo compared to inert placebo (p<0.001; 2.74±0.59 vs 0.13±0.38). **B)** Pain ratings were lower after active placebo as compared to inert placebo (p=0.002; VAS 26.2±2.0 vs 30.2±2.0), showing that the experience of side effects influenced pain relief. **C)** Most participants rated the sham condition as control (control: 93.5%; no expectation: 3.9%; fentanyl: 2.6%). The active placebo condition was believed to contain fentanyl by nearly all participants (fentanyl: 94.8%; control: 5.2%). The inert placebo condition was the most ambiguous condition: Even though participants experienced pain relief in comparison to the sham condition, more than 83% did not believe that the nasal spray contained any medication (control: 75.3%; fentanyl: 16.9%; no expectation: 7.8%). All conditions were rated significantly different from each other (all comparisons p<0.001). **D)** While their belief that side effects indicate a more effective treatment moderated the relationship between experienced side effects and their treatment expectations, the expectation about their received treatment mediated the relationship between the experienced side effects and pain relief (indirect path: -1.57(0.88); lower 95%CI: -3.83; upper 95%CI: -0.44).

When asked to rate whether they believe that a nasal spray contained fentanyl or not (Figure 2C), most participants believed that the active placebo condition contained fentanyl (94.8%). Contrary to this, the inert placebo condition was more ambiguous and only a minority of participants believed that it contained fentanyl (16.9%, p<0.001). Consistently, the confidence in their rating was larger in the active placebo condition compared to the inert placebo condition (p<0.001; Figure S2).

In a next step, we tested whether the effect of side effects depended on the participants’ expectations of having received treatment or control, as well as on their belief that side effects indicate a more potent treatment (see preregistration). We therefore conducted a moderated mediation analysis to test whether their belief that side effects indicate a more potent treatment moderated the effect of side effects on treatment expectation and if their expectation about having received fentanyl or control mediated the relationship between experienced side effects and pain relief. We observed a full moderated mediation (indirect path: -1.57(0.88); 95%CI: [-3.83 -0.44]; Figure 2D), indicating that side effects did not directly influence pain relief, but that side effects, depending on their belief about how side effects affect treatments, influence their expectations about their treatment and that these, in turn, influenced pain relief.

### Side effects lead to improved pain relief and are associated with increased rACC-PAG coupling

During the following MR session, participants were randomly allocated into an expectation group that continued to believe that pain decreases were due to fentanyl, and a no-expectation group that was explicitly debriefed that no fentanyl was present. We observed a significant interaction between expectation and side effects: The expectation group continued to show lower pain ratings after active placebo compared to inert placebo, while the no-expectation group did not show this difference any more (p=0.04; F(1,75)=4.3; Figure 3A; Figure S1). We did not observe a main effect of side effect (p=0.31) and no main effect of group (p=0.45). Side effects continued to be experienced after active placebo (p<0.001; F(1,75)=1305; Figure 3B, Figure S3) with no group difference (p=0.44) or interaction (p=0.50). This further supports the observation that pain relief is larger after experiencing a side effect and shows that this effect is modulated by the expectation of pain relief by a treatment, thereby further supporting that side effects lead to larger pain relief when treatment benefit is expected.

**Figure 3:**
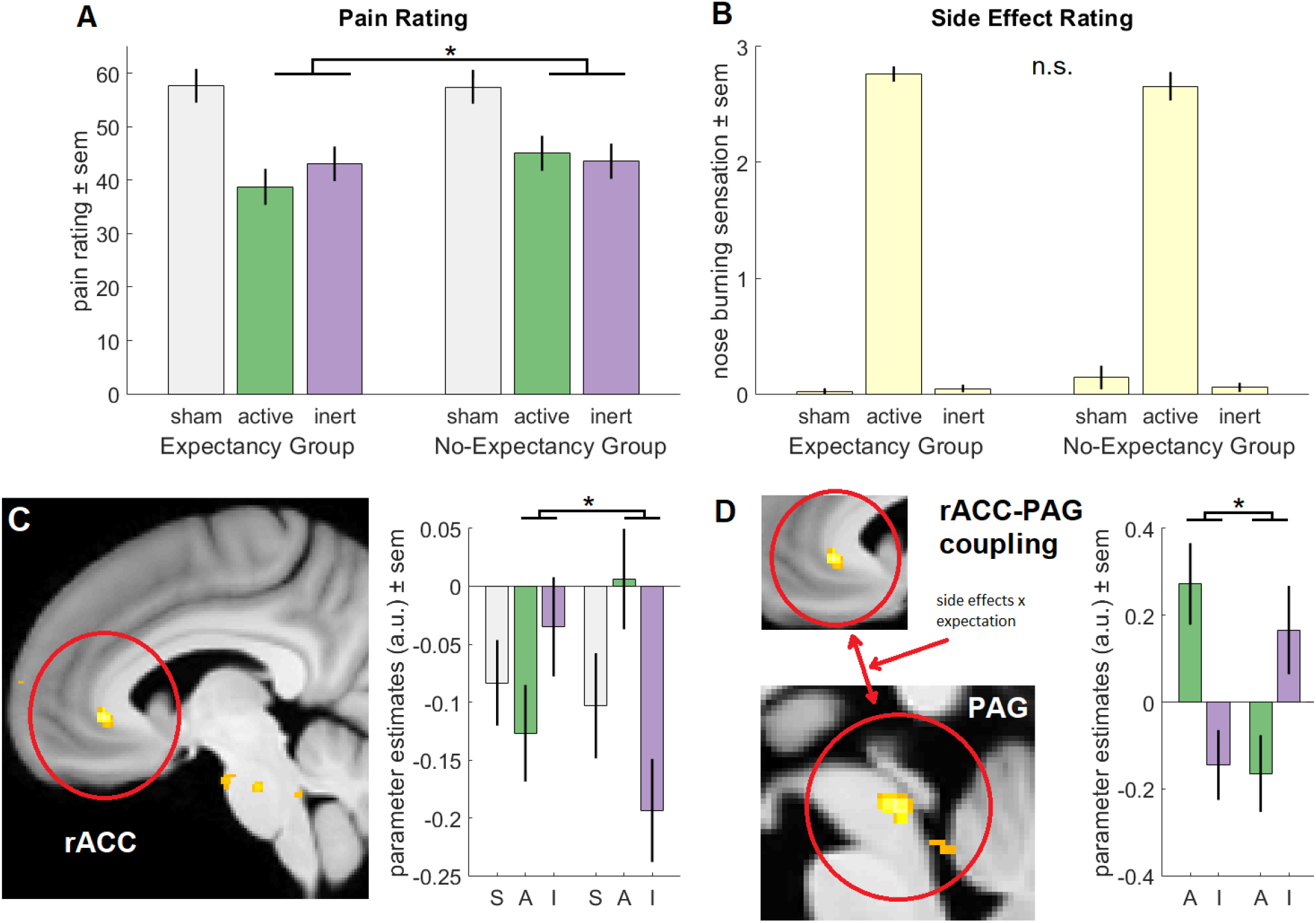
Session 2 fMRI results. Side effects lead to larger pain relief and are associated with increased rACC-PAG coupling. **A)** The expectation group continued to show lower pain ratings after active placebo compared to inert placebo, while the no-expectation group did not show this difference any more (p=0.04; VAS 38.7±3.4 vs 43.0±3.2 and VAS 45.0±3.3 vs 43.5±3.3);. **B)** Side effects continued to be experienced after active placebo with no difference between groups (p<0.001; 2.71±0.58 vs 0.05±0.22). **C)** We observed reduced BOLD signal during active placebo in the rACC (T=4.5, p=0.003; [[active_exp_ < inert_exp_] > [active_no-exp_ < inert_no-exp_]]). **D)** RACC-PAG coupling was increased during active placebo (T=4.37, p<0.001 [[active_exp_ > inert_exp_] > [active_no-exp_ > inert_no-exp_]]), indicating a stronger recruitment of the descending pain modulatory system.

We also reinvited participants to repeat the experiment 7±1 days later. As in the first session, we again observed a main effect of side effect, with lower pain ratings for the active placebo as compared to the inert placebos (p=0.004; F(1,65)=9.0). However, the group difference was not significant (p=0.15) and there was no interaction effect (p=0.93) anymore. This shows that after 7±1 days, without any further expectation manipulation, active placebo continued to lead to larger pain relief. Interestingly, the debriefed no-expectation group reestablished their expectation of treatment benefit (Figure S4).

To investigate the neural basis of how side effects interact with treatment expectations, we used fMRI to test for BOLD differences during pain stimulation. We expected that increased pain relief due to side effects might be mediated by a stronger activation of the descending pain modulatory system. Based on previous studies^14–16^, we therefore preregistered the hypothesis that the side effect by expectation interaction will be associated with a modulation of the rostral anterior cingulate cortex (rACC), the dorsolateral prefrontal cortex (DLPFC), as well as a modulation of the coupling between the rACC and the periaqueductal gray (PAG). Testing for the interaction, we observed a modulation of the rACC (T=4.5, p=0.003, [-6 33 -1.5]; Figure 3C) in the reverse contrast, indicating a reduced BOLD signal during active placebo in the expectation group, similar to previous work^15^. We did not observe a modulation of the DLPFC. We then tested if the coupling between the rACC and the PAG is increased during active vs inert placebo in the expectation group compared to the no-expectation group. We observed a significantly increased coupling (T=4.37, p<0.001, [0 -30 -12]; Figure 3D), indicating a stronger activation of the descending pain modulatory network during active placebo^17^.

## Discussion

In this study, we show that side effects can improve pain relief through the amplification of treatment expectations. We also show that expectations about treatments mediate the effect of side effects on pain relief and that this is dependent on participants’ individual beliefs of how side effects are related to treatments. Additionally, our fMRI results indicate that pain relief induced by active placebos involve a recruitment of the descending pain modulatory system.

Our data show that side effects do not directly act on the pain experience, but are mediated by treatment expectations, and are dependent on beliefs of how side effects are related to treatment effectiveness. This is in line with placebo research that shows that contextual factors primarily influence expectations that then in turn influence treatment outcome^10^. Therefore, if these mediating beliefs can be changed, treatment providers can influence how side effects affect treatment outcome.

We observed a modulation of the rACC and a stronger rACC-PAG coupling when side effects boost pain relief. Previous research with fMRI^18^, or molecular imaging^19^ has implicated this pathway in treatment expectation induced pain relief and descending pain modulation. Furthermore, rACC-PAG connectivity correlates with pain relief^15^ and blocking µ-opioid receptors with naloxone can abolish both rACC-PAG coupling and expectation induced pain relief^15^. Therefore, our results indicate that the increased expectation of treatment benefits that arise from the experience of side effects also recruits the descending pain modulatory system.

In contrast to our hypothesis, we did not observe a modulation of the DLPFC. Although the DLPFC is implicated in placebo analgesia in several studies^16^, our non-significant result with respect to the DLPFC is in line with a recent meta-analysis with individual participant data^20^, that could not confirm a significant effect in the DLPFC, possibly related to a large variability of the effect across individuals.

One additional aspect to consider is whether our effects are the result of conditioned pain modulation (CPM)^21^. In CPM a tonic painful stimulus (i.e. nasal capsaicin) renders a phasic painful stimulus (i.e. thermal pain) at a different body site less painful. However, CPM is unlikely to be relevant for our findings for two reasons: The very small dose of capsaicin inducing a mild burning sensation, as would be expected to occur as a normal treatment side effect, is unlikely to be sufficiently intense to induce CPM based on previous studies showing that a high intensity tonic stimulus is required for CPM^22^. More importantly, our observed interaction during session 2 (Figure 3A) cannot be explained by CPM as in both groups the burning sensation in the nose is identical.

Our study has also two major clinical implications. Using strategies to maximize positive treatment expectancies (i.e. placebo effects) in clinical practice can significantly improve many treatment outcomes^4,5,23^. Our data suggests that mild and benign side effects do not necessarily have to be harmful to patients and could potentially even resemble an overall benefit for treatment outcome. One could even think of artificially changing the formulation of established drug to include mild side effects to increase treatment expectations. However, while research on open label placebo^24^ provides the idea that this effect could be used without deception, great care would be necessary to avoid any unintended harm.

A simpler strategy to increase treatment benefit may lie in the framing of side effects^25^. Current expert consensus on how to inform patients about side effects emphasize the prevention of nocebo effects^26^. Studies show that optimized communication such as positive framing of side effects can indeed reduce nocebo effects^2,27^. Our mediation analysis shows that the effect of side effects on pain relief depends on beliefs that side effects are a sign of treatment effectiveness. Therefore, optimized framing of side effects as a cue that the treatment is acting and healing might not only reduce nocebo effects but also increase positive treatment expectations and placebo effects, which often lead to more beneficial treatment outcomes^4,5,28^. Our data also supports the idea that this information can be conveyed without inducing additional nocebo effects (see Supplementary Figure S3). Therefore, our study shows the psychological and neural mechanism that could be used to achieve this improvement in framing of side effects, however, it would be important to replicate the effect in a clinical sample.

Our study also has important implications for the interpretation of placebo controlled randomized clinical trials. Methodological standards such as random treatment allocation, double-blinding of patient and providers, and others have been established to achieve the assumption that nonspecific factors (i.e. placebo effects) are additive between the different experimental conditions and therefore the treatment effect can be isolated^29^. As our treatment allocation instruction was probabilistic as in most clinical trials, we can extend our findings to clinical trials and show that side effects can have a major effect on the expectations of having received treatment or having received control, consistent with previous findings^30,31^. Here, we also show that these treatment expectations then lead to differences in placebo effect. Our data supports that side effects can differentially influence placebo effects when side effect occurrence differs between experimental conditions, as is often the case in clinical trials^32,33^. The validity of clinical trials depends on the additivity of nonspecific factors (i.e. placebo effect). Our data shows that this is not the case if side effects differ between treatment and control arm and therefore question the validity of the additivity assumption in these cases. As a consequence, clinical trials could overestimate the effect of a drug if side effects increase placebo effects in the treatment condition. Active placebos in the control arm^9^ or other innovative research designs^34^ could counteract this confounds and improve clinical trial validity. Taken together, our data shows the significant influence of side effects on treatment expectations and placebo effects. Taking these effects into account could improve clinical practice as well as clinical trials.

## Data availability statement

All non-identifiable data produced in the present study are available upon reasonable request to the authors.

## Acknowledgements

We would like to thank Daniel Rathmann, Nora Kösters and Justus Lübbemeier for their dedicated support during data collection. We also thank Björn Horing for administrative support during funding acquisition as well as Ulrike Bingel and Winfried Rief for comments on an earlier version of this manuscript.

## Author contributions

Conceptualization, methodology, investigation, data curation, formal analysis, visualization, project administration, writing – original draft, L.S.; Methodology, investigation, validation, writing – review and editing T.F.; Conceptualization, methodology, resources, funding acquisition, supervision, validation, project administration, writing – review and editing, C.B.

## References

1. Petrie KJ, Rief W. Psychobiological Mechanisms of Placebo and Nocebo Effects: Pathways to Improve Treatments and Reduce Side Effects. Annu Rev Psychol. 2019;70:599–625. doi:10.1146/annurev-psych-010418-102907

2. Colloca L, Miller FG. The Nocebo Effect and Its Relevance for Clinical Practice. Psychosom Med. 2011;73(7):598. doi:10.1097/PSY.0b013e3182294a50

3. Berna C, Kirsch I, Zion SR, et al. Side effects can enhance treatment response through expectancy effects: an experimental analgesic randomized controlled trial. Pain. 2017;158(6):1014–1020. doi:10.1097/j.pain.0000000000000870

4. Benedetti F. Placebo and the New Physiology of the Doctor-Patient Relationship. Physiol Rev. 2013;93(3):1207–1246. doi:10.1152/physrev.00043.2012

5. Kaptchuk TJ, Miller FG. Placebo Effects in Medicine. N Engl J Med. 2015;373(1):8–9. doi:10.1056/NEJMp1504023

6. Sanders AE, Slade GD, Fillingim RB, Ohrbach R, Arbes SJ Jr, Tchivileva IE. Effect of Treatment Expectation on Placebo Response and Analgesic Efficacy: A Secondary Aim in a Randomized Clinical Trial. JAMA Netw Open. 2020;3(4):e202907. doi:10.1001/jamanetworkopen.2020.2907

7. Rief W, Glombiewski JA. The hidden effects of blinded, placebo-controlled randomized trials: An experimental investigation. PAIN. 2012;153(12):2473–2477. doi:10.1016/j.pain.2012.09.007

8. Linde K, Atmann O, Meissner K, et al. How often do general practitioners use placebos and non-specific interventions? Systematic review and meta-analysis of surveys. PLOS ONE. 2018;13(8):e0202211. doi:10.1371/journal.pone.0202211

9. Jensen JS, Bielefeldt AØ, Hróbjartsson A. Active placebo control groups of pharmacological interventions were rarely used but merited serious consideration: a methodological overview. J Clin Epidemiol. 2017;87:35–46. doi:10.1016/j.jclinepi.2017.03.001

10. Montgomery GH, Kirsch I. Classical conditioning and the placebo effect. Pain. 1997;72(1– 2):107–113. doi:10.1016/S0304-3959(97)00016-X

11. Atlas LY, Wager TD. A Meta-analysis of Brain Mechanisms of Placebo Analgesia: Consistent Findings and Unanswered Questions. In: Benedetti F, Enck P, Frisaldi E, Schedlowski M, eds. Placebo. Springer Berlin Heidelberg; 2014:37-69.

12. Linnman C, Moulton EA, Barmettler G, Becerra L, Borsook D. Neuroimaging of the periaqueductal gray: State of the field. NeuroImage. 2012;60(1):505–522. doi:10.1016/j.neuroimage.2011.11.095

13. Friston KJ, Buechel C, Fink GR, Morris J, Rolls E, Dolan RJ. Psychophysiological and Modulatory Interactions in Neuroimaging. NeuroImage. 1997;6(3):218–229. doi:10.1006/nimg.1997.0291

14. Bingel U, Lorenz J, Schoell E, Weiller C, Büchel C. Mechanisms of placebo analgesia: rACC recruitment of a subcortical antinociceptive network. Pain. 2006;120(1–2):8–15. doi:10.1016/j.pain.2005.08.027

15. Eippert F, Bingel U, Schoell ED, et al. Activation of the opioidergic descending pain control system underlies placebo analgesia. Neuron. 2009;63(4):533.

16. Wager TD, Atlas LY. The neuroscience of placebo effects: connecting context, learning and health. Nat Rev Neurosci. 2015;16(7):403–418. doi:10.1038/nrn3976

17. Bingel U, Tracey I. Imaging CNS Modulation of Pain in Humans. Physiology. 2008;23(6):371–380. doi:10.1152/physiol.00024.2008

18. Wager TD, Rilling JK, Smith EE, et al. Placebo-Induced Changes in fMRI in the Anticipation and Experience of Pain. Science. 2004;303(5661):1162-1167. doi:10.1126/science.1093065

19. Pecina M, Azhar H, Love TM, et al. Personality Trait Predictors of Placebo Analgesia and Neurobiological Correlates. Neuropsychopharmacology. 2013;38(4):639–646. doi:10.1038/npp.2012.227

20. Zunhammer M, Spisák T, Wager TD, Bingel U. Meta-analysis of neural systems underlying placebo analgesia from individual participant fMRI data. Nat Commun. 2021;12:1391. doi:10.1038/s41467-021-21179-3

21. Petersen KK, McPhee ME, Hoegh MS, Graven-Nielsen T. Assessment of conditioned pain modulation in healthy participants and patients with chronic pain: manifestations and implications for pain progression. Curr Opin Support Palliat Care. 2019;13(2):99–106. doi:10.1097/SPC.0000000000000419

22. Granot M, Weissman-Fogel I, Crispel Y, et al. Determinants of endogenous analgesia magnitude in a diffuse noxious inhibitory control (DNIC) paradigm: Do conditioning stimulus painfulness, gender and personality variables matter? PAIN®. 2008;136(1):142–149. doi:10.1016/j.pain.2007.06.029

23. Colloca L, Barsky AJ. Placebo and Nocebo Effects. N Engl J Med. 2020;382(6):554-561. doi:10.1056/NEJMra1907805

24. Kaptchuk TJ, Miller FG. Open label placebo: can honestly prescribed placebos evoke meaningful therapeutic benefits? BMJ. 2018;363:k3889. doi:10.1136/bmj.k3889

25. Leibowitz KA, Howe LC, Crum AJ. Changing mindsets about side effects. BMJ Open. 2021;11(2):e040134. doi:10.1136/bmjopen-2020-040134

26. Evers AWM, Colloca L, Blease C, et al. Implications of Placebo and Nocebo Effects for Clinical Practice: Expert Consensus. Psychother Psychosom. 2018;87(4):204–210. doi:10.1159/000490354

27. Manaï M, van Middendorp H, Veldhuijzen DS, Huizinga TWJ, Evers AWM. How to prevent, minimize, or extinguish nocebo effects in pain: a narrative review on mechanisms, predictors, and interventions. PAIN Rep. 2019;4(3):e699. doi:10.1097/PR9.0000000000000699

28. Bingel U. Placebo 2.0: the impact of expectations on analgesic treatment outcome. PAIN. 2020;161:S48. doi:10.1097/j.pain.0000000000001981

29. Sibbald B, Roland M. Understanding controlled trials: Why are randomised controlled trials important? BMJ. 1998;316(7126):201. doi:10.1136/bmj.316.7126.201

30. Turner JA, Jensen MP, Warms CA, Cardenas DD. Blinding effectiveness and association of pretreatment expectations with pain improvement in a double-blind randomized controlled trial. PAIN. 2002;99(1):91–99. doi:10.1016/S0304-3959(02)00060-X

31. Basoglu M, Marks I, Livanou M, Swinson R. Double-blindness Procedures, Rater Blindness, and Ratings of Outcome: Observations From a Controlled Trial. Arch Gen Psychiatry. 1997;54(8):744–748. doi:10.1001/archpsyc.1997.01830200078011

32. Bell RF, Eccleston C, Kalso EA. Ketamine as an adjuvant to opioids for cancer pain. Cochrane Database Syst Rev. 2017;6(6):CD003351. doi:10.1002/14651858.CD003351.pub3

33. Els C, Jackson TD, Kunyk D, et al. Adverse events associated with medium- and long-term use of opioids for chronic non-cancer pain: an overview of Cochrane Reviews. Cochrane Database Syst Rev. 2017;2017(10):CD012509. doi:10.1002/14651858.CD012509.pub2

34. Colagiuri B. Participant expectancies in double-blind randomized placebo-controlled trials: potential limitations to trial validity. Clin Trials. 2010;7(3):246–255. doi:10.1177/1740774510367916

