## Supplementary Material for "How side effects can improve treatment efficacy: a randomized trial"

**Clinical Trials Registration:** https://drks.de/search/de/trial/DRKS00026648

### Supplementary Methods

#### Participants

Participants were confirmed to be healthy through an in-person interview with a medical doctor during the initial visit. Inclusion criteria included age between 18-45 and BMI 18-30. Exclusion criteria included somatic or psychiatric conditions, regular intake of medications (excl. hormonal contraception, allergy medications, thyroid medication), recent (<24h) intake of pain medication, drug abuse, pregnancy or breast feeding, skin condition or damage on forearms, nasal condition or damage, allergy against pain medications, as well as any conditions that might affect MRI safety. Participants performed a drug test during the initial visit and were ineligible to participate in case of a positive result. Participants were recruited using flyers and internet advertisements and were compensated for their participation. Data collection was performed between October 2021 and June 2022 at the University Medical Center Hamburg-Eppendorf.

#### Manipulation Check

After each experiment, a structured interview was used as a data-of-interest independent measurement to determine if participants were convinced by the cover story and if they actually believed that there was a possibility for them to receive fentanyl. This was done to ensure validity as a successful cover story was necessary to study treatment expectation effects. After the structured interview, two separate scientists (T.F. and L.S.) read participant’s statements independently and rated if the cover story was believed successfully. In case of disagreement, a third scientist (C.B.) read the statements and gave the deciding vote. This was done for all participants independently to avoid inducing group differences.

#### 7±1 days follow-up visit

All participants were invited for a 7±1 day follow-up experiment, however some participants declined the follow-up experiment or were unable to attend. During the follow-up visit, participants completed the same paradigm as before, with pain corresponding to VAS60 in the second and third run (Supplementary Figure 2). 67 participants were included in the data analysis for the follow-up experiment (36 placebo group (age: 24.5±3.9 [20-38], 11 male); 31 control group (age: 24.9±5.6 [18-43], 12 male)).

#### Data acquisition

Stimulus presentation and behavioral data collection was carried out using Psychtoolbox in MATLAB (MathWorks, Natick, MA, USA). FMRI data were acquired on a 3-Tesla system (Magnetom Prisma, Siemens, Erlangen, Germany) equipped with a 64-channel head coil. To measure BOLD responses, a T2*-weighted 2D multiband gradient echo planar imaging sequence was used (TR: 1.9s; TE: 26ms; field of view: 225 $mm^2$; multiband acceleration: 3). Each volume consisted of 78 transversal slices with a voxel size of 1.5x1.5x1.5$mm^3$. High resolution anatomical T1-scans were acquired using an MPRAGE sequence with a voxel size of 1x1x1$mm^3$.

### Supplementary Figures

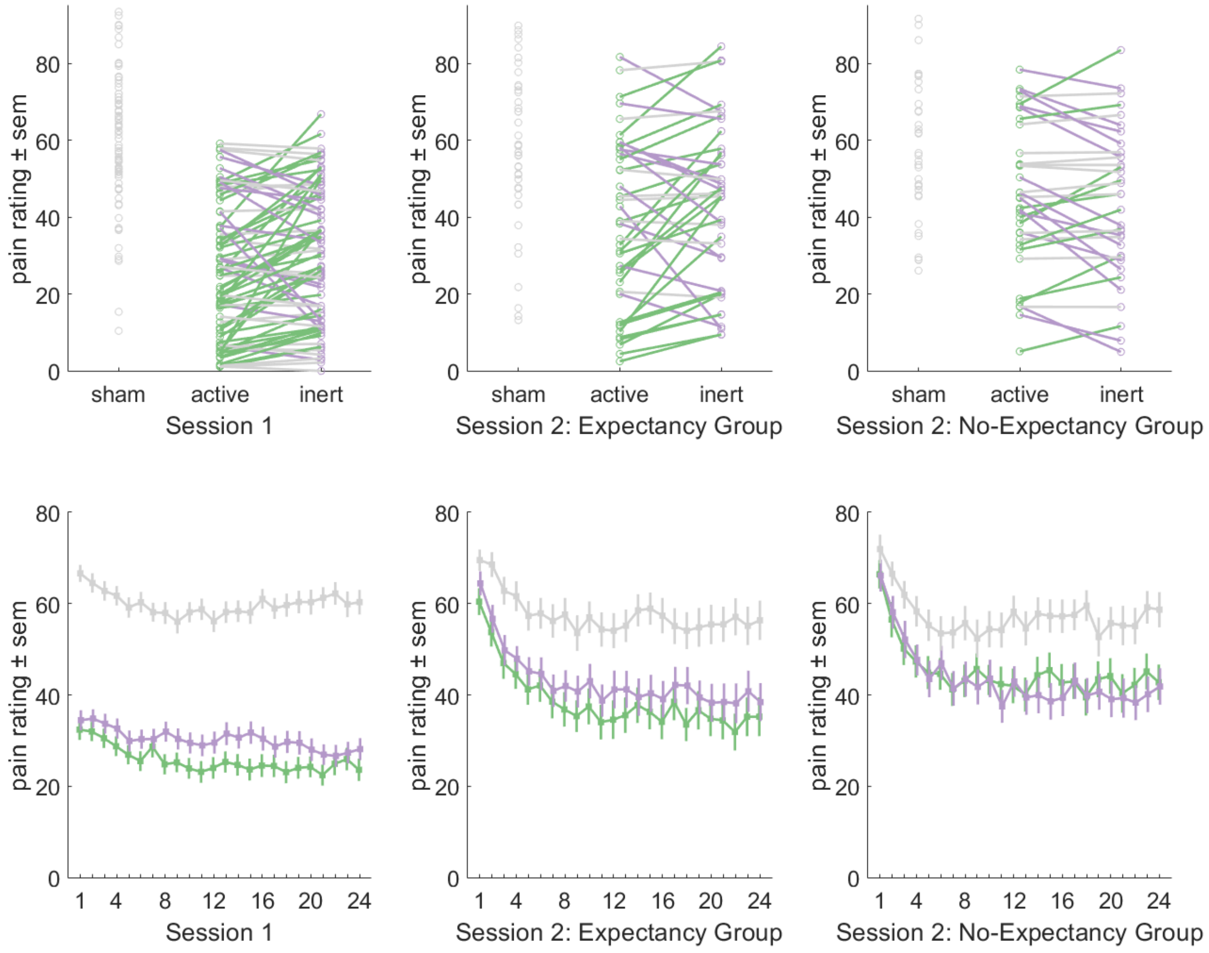

**Supplementary Figure S1: Individual pain ratings and pain ratings over time**

During session 1, pain ratings were lower after active placebo as compared to inert placebo. During session 2, the expectation group continued to show lower pain ratings after active placebo compared to inert placebo, while the no-expectation group did not show this difference any more. Circles and lines indicate individual values and differences. A green line indicates lower pain during active placebo (<-3), a grey line indicates no difference (-3<x<3) and a purple line indicates a higher pain during active placebo (>3).

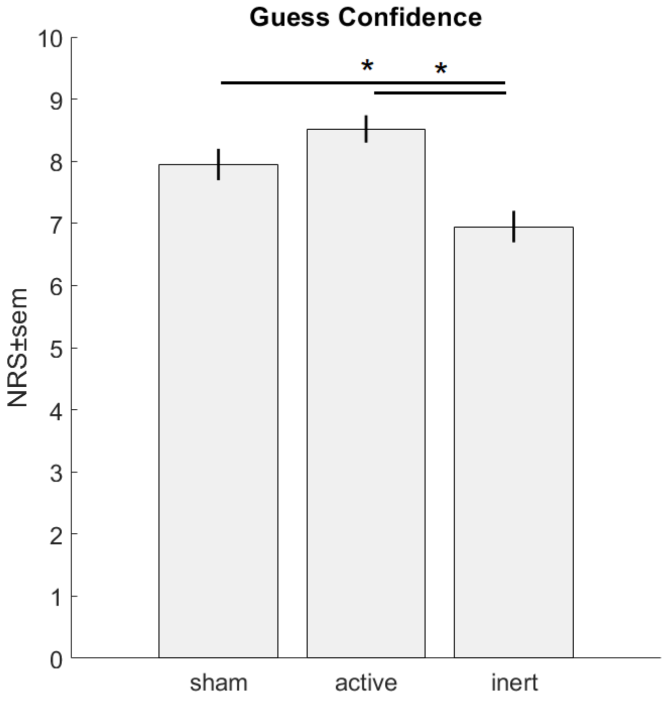

**Supplementary Figure S2: Guess confidence for the treatment expectation**

The confidence ratings were generally high (sham: 8.0±0.25/10; active: 8.5±0.22/10; inert 7.0±0.26/10). Confidence ratings were lower for the inert condition as compared to the sham and active condition ($p = 0.003$ and $p < 0.001$, Paired-T-tests, Bonferroni-corrected). The comparison between sham and active failed to reach statistical significance after correction for multiple testing ($p = 0.036_{uncorrected}$).

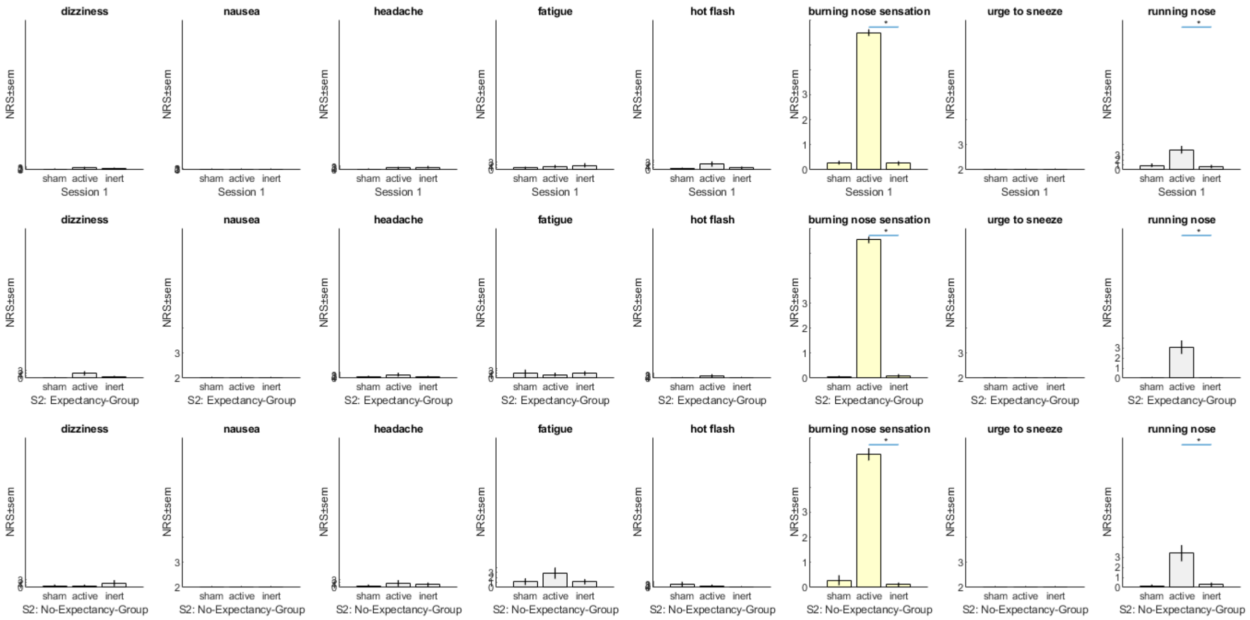

#### Supplementary Figure S3: all side effect reports

Participants reported no additional side effects that were not related to the capsaicin application in the nose in the active placebo condition compared to the inert placebo condition (all p>0.05). Participants reported more burning sensation in the nose (all p<0.001; targeted side effect; yellow) and more runny nose (all p<0.05; grey) in the active compared to the inert placebo condition.

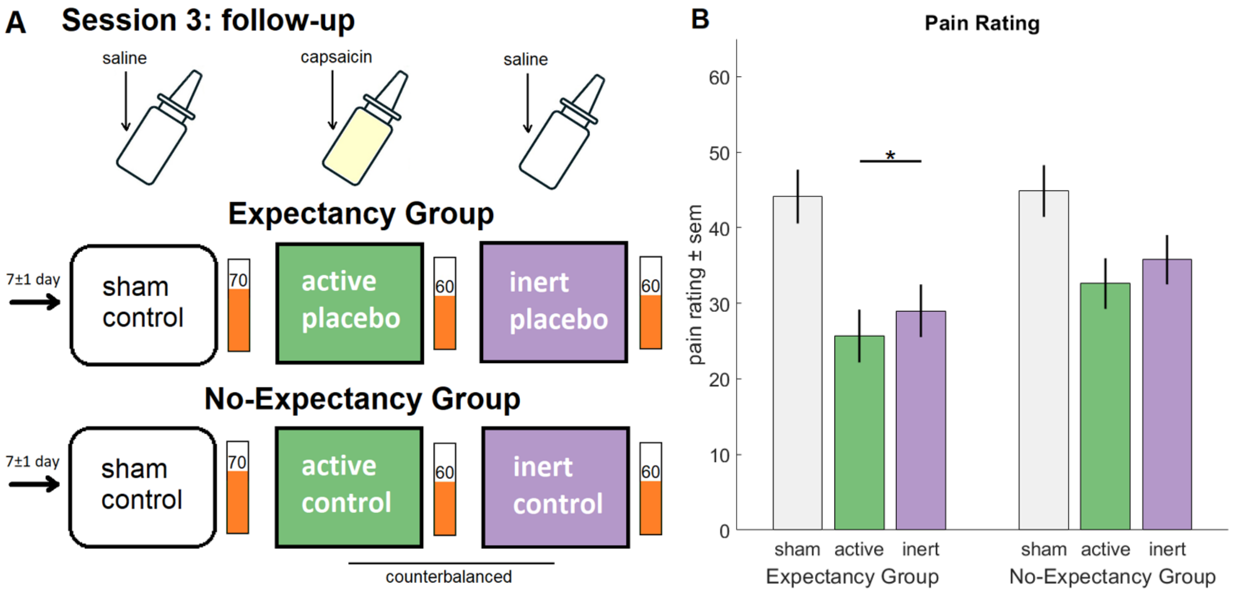

**Supplementary Figure S4: 7±1 day follow up investigation**

**A)** During the 7±1 day follow up, 36 participants of the expectation group and 31 participants of the no-expectation group received the same 3 nasal sprays. **B)** We observed a main effect of side effect, with lower pain ratings for the active placebo as compared to the inert placebos ($p = 0.004$; $F(1,65)=9.0$; VAS 28.9±2.5 vs 32.1±2.5). The group difference ($p = 0.15$) and the interaction ($p = 0.93$) was not significant. Subsequent contrasts showed a significant difference in the expectation group ($p = 0.03$) but only marginal statistical significance in the no-expectation group ($p = 0.05$). Therefore, side effects boosted treatment even in the group that was explicitly told that the nasal spray did not contain any fentanyl. In contrast to the fMRI experiment which was conducted directly after debriefing, after 7±1 days and without any further expectancy manipulation, active placebo again lead to larger placebo effects, suggesting a reestablished belief of additional benefit through side effects, potentially through contextual conditioning[1]. This data is in line with research on open-label placebos suggesting that placebo treatments can be effective even if participants know that they are only receiving a placebo treatment[2,3] and underscore the profound effect of experiencing side effects on placebo effects.

#### References

1. Colloca L, Miller FG. How placebo responses are formed: a learning perspective. Philos Trans R Soc Lond B Biol Sci 2011;366(1572):1859–69.

2. Kaptchuk TJ, Miller FG. Open label placebo: can honestly prescribed placebos evoke meaningful therapeutic benefits? BMJ 2018;363:k3889.

3. Charlesworth JEG, Petkovic G, Kelley JM, et al. Effects of placebos without deception compared with no treatment: A systematic review and meta-analysis. J Evid-Based Med 2017;10(2):97–107.
